# Bridging the Heterogeneity of Myasthenia Gravis Severity Scores for Digital Twin Development

**DOI:** 10.1101/2025.06.13.25329566

**Authors:** Marc Garbey, Quentin Lesport, Henry J. Kaminski

**Affiliations:** Care Constitution Corp - Houston, TX, USA; LaSIE UMR-CNRS 7356 University of La Rochelle, France; Department of Neurology & Rehabilitation Medicine, George Washington University, Washington DC, USA

**Keywords:** precision medicine, digital twin, myasthenia gravis, ptosis, diplopia, muscular weakness, computer vision, eyes tracking, neurological disease, clinical trial, virtual population

## Abstract

Myasthenia gravis (MG) is a rare autoimmune neuromuscular disease. Clinical trials with rigorously collected data, especially for rare diseases, provide opportunities for mathematical modeling of patient outcomes over time; however, building a larger data set from multiple trials faces the challenge of harmonization of outcome measures. To accurately model MG and predict individual patient trajectories, one requires integrating three primary data types: (i) Laboratory and medication data, (ii) Electronic Health Record (EHR) data (e.g., age, sex, years since diagnosis, BMI), (iii) Disease severity scores.

Among these, MG severity scores are crucial for measuring disease progression from the patient’s and clinical evaluator’s perspectives. However, clinical studies often employ various scoring systems (e.g., ADL, QMG, MG-CE, MGQOL-15), making it challenging to determine the most reliable measure. In this study, we investigate the relationships among clinical outcome measures across multiple clinical studies. Our objective is to develop a robust “Myasthenia Gravis Portrait” that can be applied across diverse clinical studies. This standardized portrait will facilitate the creation of a virtual population of digital twins, enabling the application of machine learning techniques to a larger patient population.

## Introduction

Myasthenia gravis (MG) is a rare, autoimmune, neuromuscular disorder caused primarily by antibodies directed towards the acetylcholine receptor (AChR) on the post-synaptic surface of the neuromuscular junction, with fluctuating weaknessmanifest as disabling visual symptoms to life-threatening respiratory failure [1]. There is significant unmet need with poor adverse effect profiles of therapies, wide variation of responses, and a third of patients being treatment-resistant with recent FDA-approved medications having a poorer than expected benefit in common clinical practice.

Validated clinical outcome measures used to assess (MG patients have been established some being based on symptoms reported by the patient, others by clinical examination, and some are hybrid. The most commonly used are:

- Quantitative Myasthenia Gravis (QMG) Score: The QMG was is based on physical examination of sentinel muscle groups compromised in MG [2, 3].
- Myasthenia Gravis Composite (MGC) Score: This score is based on physician examination and patient reported impairment in performance of specific activities [4, 5].
- Myasthenia Gravis Activities of Daily Living (MG-ADL) Scale: This patient-reported outcome measure assesses MG symptoms and functional activities related to daily living. The score is the primary outcome measure used by the FDA for drug approval [6, 7].
- Myasthenia Gravis Fatigue Scale (MG-FS): This patient self-reported scale assesses fatigue in MG patients, which is a common complaint [8].

Unfortunately, those scores might be computed using slightly different version than the MG-FS scale such as the MG-QOL15 for the fatigue score as defined in [9]. The new MG-Core Exam (CE) score was developed to adapt the neuromuscular examination for performance for telemedicine conditions [10]. A common feature to all these scores is the heuristic nature of the data acquisition and potential bias due to the method of reporting and/or observation. The QMG and ADL score have essential differences: The QMG provides a quantitative measure of muscle strength at a single point in time performed by a trained investigator or research coordinator. The MG-ADL provides a measure of the patient’s perception in performing activities in the previous two weeks of evaluation [11]. t The MGC score is a combination of both ADL and QMG scores, in an attempt to provide a comprehensive assessment of MG severity, combining physician-assessed examinations (evaluates ocular, neck, and proximal limb muscles) and patient-reported outcomes (assesses speech, chewing, swallowing, and respiratory function). Our goal is to provide a clinical score portrait that goes beyond this simple dichotomy and support a digital twin construction from multiple disparate clinical studies.

In this paper, we use data sets from three clinical studies: the ADAPT-TeleMG data set [10, 12, 13], MGTX trial [14, 15], Beat-MG trial [16] to build a patient clinical portrait that can be used as a basis in a common construction of a Virtual Population of Digital Twins (VPTD) [17].

Each of these studies had different end points and goals. Therefore, it is not surprising that their population distributions obey different criteria. Clinical studies do not use the same standard score. Since MG is a rare disease and clinical trial recruitments range from fewer than 50 to 200, it will be essential to combine multiple studies to design a digital twin of the MG patient that would have the statistical power to support disease prediction. Consequently, our goal is to consolidate whatever score is available from a study into a single clinical portrait of the MG patient.

First, we will look within each score at its components and their relationship to each other. Mathieu et al. [18] provides a thorough statistical analysis of this problem for the MG-ADL score. Second, we will look at the potential correlation across outcome measures for corresponding muscle group evaluations. Our focus is to build stochastic mapping components from one score to another, to build a “portrait “of the subject and fill potential gaps across clinical trials. A patient’s MG score portrait should be a comprehensive assessment of the disease’s severity and impact on their daily life. We construct a consolidated score from disparate incomplete ones using simple techniques including clustering and descriptive statistical methods. Together, these components paint a clinical picture of a patient’s disease and would assist healthcare providers in tailoring a treatment plan, assess treatment effectiveness, and provide prognostic information. In our previous study [19], we used the MGTX trial to develop a modeling framework aimed at enhancing the efficiency and predictive power for clinical trials, using MG as a representative example of a rare disease. This study intends to allow the generalization of this work to a larger number of clinical trials by providing a much broader base of patient data.

## Methods

### Data Sets

- ADAPT-teleMG (NCT05917184): We utilized a bank of 54 subjects with MG each having two telemedicine examinations by neuromuscular experts with recording of the MG-CE score, the MG-ADL, MG-Composite Virtual, MG-CE and Neuro_QOL15 score [9]. The total number of records was 104 in each category. A critical review of the human factors influencing the examination scores is provided in [13].
- Beat MG (B-cell depletion in MG) [16]: The Phase 2 trial evaluated the use of rituximab, a medication that depletes B cells in patients with MG who were not adequately controlled with standard treatments. 52 subjects were followed over a period of approximatively one year. The primary clinical outcome measure was based on prednisone dose and MGC score. The total number of ADL and QMG scores accumulated over all patient and over time was 728.
- MGTX (Thymectomy Trial in Non-Thymomatous MG Patients Receiving Prednisone Therapy) [14, 15]: This 3 years trial investigated the effectiveness of thymectomy plus prednisone versus prednisone alone in patients with generalized MG. Subjects were 18 to 65 years and were AChR-antibody positive. Subjects doing poorly had escalation of prednisone, and for severe worsening, treated with rescue therapies of IVIg or plasma exchange. One hundred and twenty-three subjects followed for 36 months. The primary outcome was the average alternate-day prednisone dose: This measured the amount of prednisone (a corticosteroid medication used to suppress the immune system) needed to control MG symptoms. The ADL and QMG score were to be done at month M0, M3, M4, M6, M9, M12, M15, …M36.The total number of ADL sand QMG scores accumulated over all patients over time was 2061, with some missing entries for each of the outcome measures

### Concept

Our construction of a patient portrait is based on establishing stochastic maps between individual test score values of the various outcome measures, which assess the same group of muscles. This construction should gather the accumulated knowledge of all clinical trial data sets. This mapping should be done in the context of possible existing correlation between individual test scores within a score.

For example, the construction of the two stochastic maps that project the ADL double vision/Eyelid Drop score to the QMG ptosis/diplopia score and vice versa is based on all the clinical data sets available that is about 2800 entries combining MGTX and BeatMG. It can be generated either by enumeration counts or machine learning.

The mathematical formulation of the clinical score portrait of a patient gathering QMG and ADL information will come as a combination of discrete values from all available scores and probability distribution of score values when a score is missing. Let us assume for example that a patient received a standard QMG score but the corresponding ADL score for the visit is missing. Using a stochastic map from QMG to ADL, we can propose to provide the ADL missing record with the probability distribution corresponding to the ADL score. The table below shows an example for an individual score, ptosis with provided QMG score but missing ADL score:

**Table 1.** Example of score values using a combination of measured outcome for QMG and estimated outcome for ADL.

| Score Value | 0 | 1 | 2 | 3 |
| --- | --- | --- | --- | --- |
| QMG | 0 | 1 | 0 | 0 |
| ADL | 0.6 | 0.3 | 0.1 | 0.1 |

The more precise is the stochastic map, the better is the estimate of the missing score As we build the stochastic map from all available data, we would continue to improve upon the estimate.

In situations where two test individual scores denoted A and B have demonstrated some level of correlation, we may use stochastic maps combining both individual test scores to improve the estimate.

A “two dimensional” score estimate would take the format of the following matrix,

**Table 2.** The output of the stochastic map provides a probability distribution for the couple of possible score evaluations (score A, score B)

| Test score A / Test Score B | 0 | 1 | 2 | 3 |
| --- | --- | --- | --- | --- |
| 0 | 0.1 | 0.1 | 0 | 0 |
| 1 | 0.5 | 0.2 | 0 | 0 |
| 2 | 0. | 0. | 0.1 | 0. |
| 3 | 0. | 0. | 0. | 0. |

The same concept applies to more than two individual test scores depending on the level of correlation of inter-individual test scores.

If the score has been given twice or more by different clinicians, based on evaluation of video records of examinations of the same patient as was the case in the ADAPT clinical study, we may have different scores from clinicians. We replace the multiple entries of the score by a probability estimate assuming either the same weight to each clinician, or possibly increase the trust on one of the evaluators if there were objective criteria to do so [10, 12, 13].

Assuming that all the individual components of the QoL test score are strongly corelated as opposed to the ADL and QMG scores, as we will demonstrate later, the construction of the stochastic map between the QoL score and ADL or QMG is more complicated.

It is no surprise that the answer of a patient to a generic question such as “I am frustrated by my MG” could be related to “I have trouble eating because of MG”. This is true also for “I have trouble with my eyes because of my MG (e.g. double vision)” and “I have limited my social activity because of my MG” to take a few examples.

We use clustering to transform individual test scores into few clusters that illustrate the patient fatigue pattern from QoL and map directly this pattern to the total score for ADL and QMG or possibly its components.

Overall, we can complete the individual score of any patient enrolled in one of the clinical trials with a uniform structure for the score portrait that combine elements of the ADL, QMG and QoL scores for example and can serve as a basis to predict patient trajectories as reported in [19]. We can also refine our strategy in assembling the score portrait, by using our predictability index associated to the data set to obtain the best weighted possible combination of scores, in the format for example,

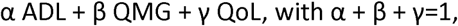

or at the level of individual test scores within ADL or QMG as shown in [19].

Since the patient portrait is a probability distribution, we can generate readily a virtual population [20, 21] using a Montecarlo simulation and minimum assumption. This is indeed a resampling method that does not add new diverse patient profile and works without any modeling assumption on the patient response to treatment. We will apply our patient trajectory scheme to this virtual population to refine our previous results on treatment success, azathioprine doses in particular, revealing potential new associations with age and rescue therapy [22].

To summarize, our construction follows the algorithm described next along with its rational.

### Algorithm

- Step 1: each score corresponding to muscle weakness symptoms is assembled from a battery of individual test score generally focusing on a specific group of muscle:

- Compute the correlation matrix between individual test scores of the same score for each data set and check if correlation between individual score is weak. The concept of weak correlation is indeed somewhat fuzzy and will be define in the result section. If correlation between individual test score is not weak regroup them as a vector.
- Compute the distribution of individual test scores for each score and data set.
- Step 2: Review the fatigue test score with a similar method:

- show that the correlation between individual scores is strong.
- Extract a set of independent patterns with weak correlation using a clustering algorithm.
- We note that of all three data sets we used, only the MGnet data set has a QoL score, more precisely the Neuro-Qol 15 version.
- Step 3: Establish potential correlation between score test across scores (ref step 1): putting aside the fatigue test, revisit the graph of correlation inter-score see Table 3 & 4: Leg strength/double vision/Eyelid Drop is the smallest common denominator between all first three scores. All other test score might be re-group by domains: talking, chewing, swallowing, count to 50 (bulbar domain); breathing, single breath count (respiratory domain); impairment of ability to brush teeth or comb hair, impairment of ability to rise from a chair (limb weakness domain); and double vision, eyelid droop, eye closure (ocular domain).

**Table 3:** List of individual test scores for each outcome measure.

| Score | List of Individual Test Score |
| --- | --- |
| MG-ADL | Talking/Chewing/Swallowing/Breathing/Brush Teeth/Sit to Stand/double vision/Eyelid Drop |
| MG-CE | Ptosis/Diplopia/Cheek Puff/Tong to Cheek/Count to 50/Arm sFatigue/Single Breath/Sit to Stand |
| QMG | Left Leg/Right Leg/Head/Left Grip/Right Grip/Vital/Left Arm/Right Arm/Speech/Swallow/Facial/Ptosis/Vision |

- Step 4: Extend that correlation matrix to the QoL patterns (ref step 2) of QoL Scores: Frustration/Ocular Impairment/Social Activity/Hobbies Activity/Family Duties/Making Plan/Work ability/Speaking/Lost of Autonomy/Depression/Walking/Moving to place/Overwhelmed/Personal Grooming.
- Step 5: generate a virtual population with a Monte Carlo simulation starting from the stochastic portrait of each patient data set entered in the data base.

**Table 4:** Organization of Symptoms from Scores by Group.

| Group 1 | Group 2 | Group 3 | Group 4 | Group 5 | Group 6 |
| --- | --- | --- | --- | --- | --- |
| Ptosis | Arm Strength | Breath | Sit to Stand | Talking | Chewing |
| Diplopia | Arm Extension | Single Breath Count | Leg Strength | Count to 50 | Swallowing |
| Double Vision | Brush Teeth |  |  |  | Tong to Cheek |
| Eye lid drop | Grip |  |  |  | Talking |
|  |  |  |  |  | Facial |

Once this construction is done, we can study patient score trajectories during the clinical trial and use the methodology of [16] to start to transform that data set into a predictive model of the clinical trial.

## Result

We appreciate that the self-reported scores like ADL or QoL have a level of uncertainty [23] due to the subjectivity of the subject’s perception, mood, and emotional state. Further, symptoms fluctuate significantly and the patient may develop coping strategies, which under-estimate ther the severity of symptoms. Similarly, clinical examination with QMG and MG-CE score are not perfect either, and may have poor reproducibility in some categories - see Figure1 and [13]. Eventually, score values in clinical examination score might be more robust and accurate with digital health technique than with clinician observations [10, 12].For these reasons. we expect then a significant level of noise and bias in our data set that could lower the robustness of the analysis.

**Figure 1:**
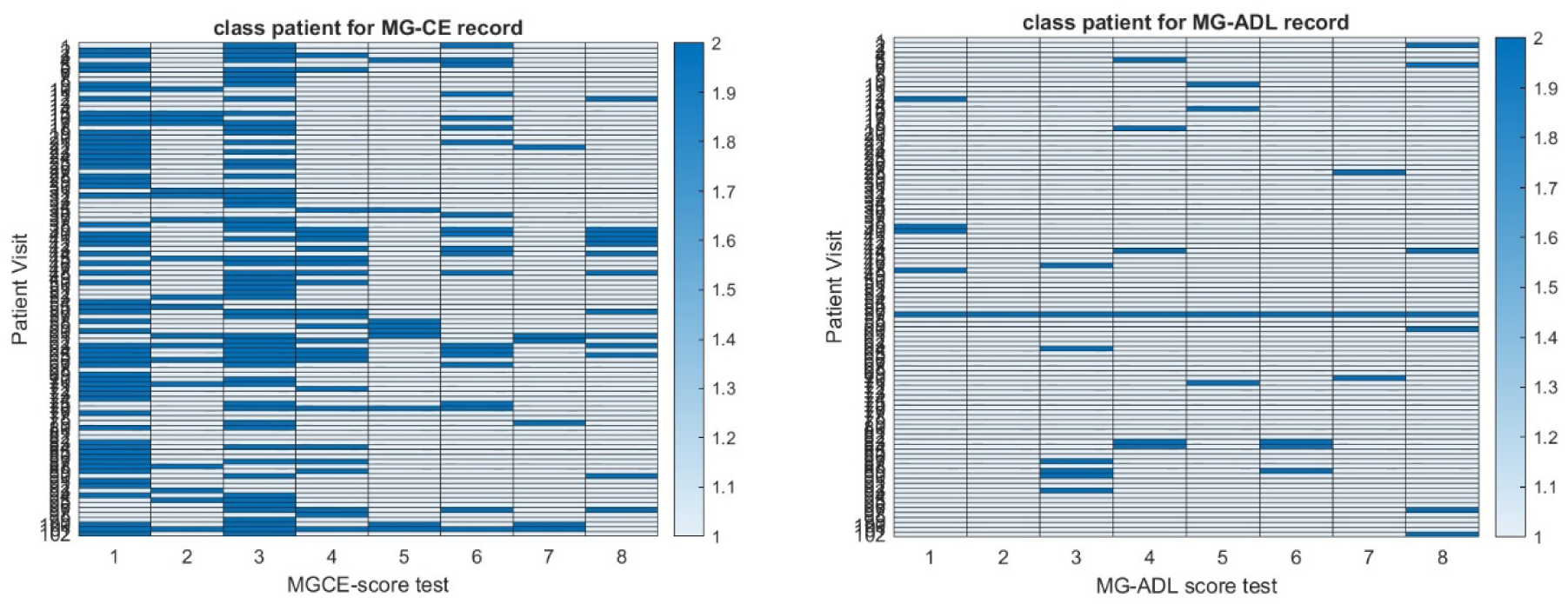
Heat map of the Reproducibility of MG-CE score (left panel) and MG-ADL score (right panel) in the ADAPT-MG study as defined in [11].

We assessed each clinical data set for the distribution of patient score and the percentage of the population with positive symptoms using ADL for each score test – see Figure 2. We generate ρ a generic correlation coefficient and its p-values denoted p. All three distributions are distinct but similar. The average scores between the three data sets have the best correlation with ρ>0.85, p<=0.007. It should be appreciated that while the number of patients is roughly of the same order for each data set, the number of registered scores we can consider as independent for our purpose study is not. In fact scores are determined at different time points of the clinical study and can be considered at independent variables, since we are only interested in correlation across scores.

**Figure 2:**
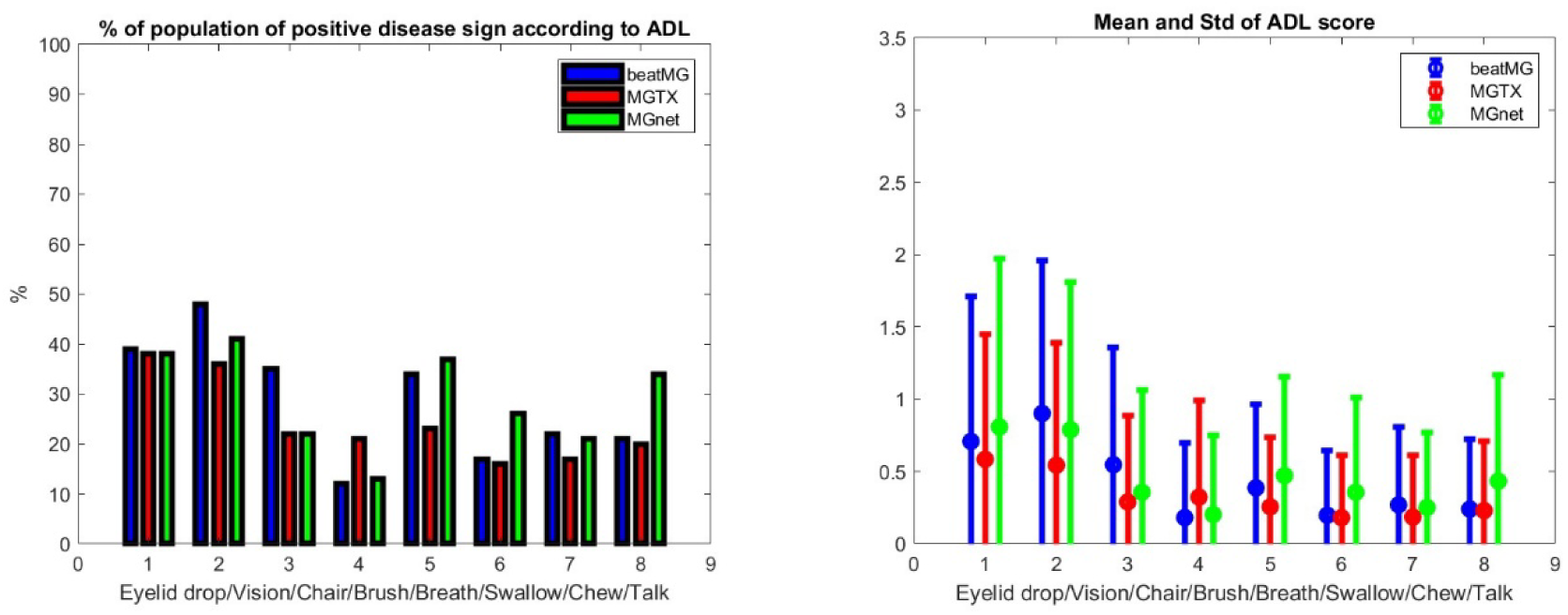
Population Characteristics of the beatMG, MGTX and ADAPT-MG clinical studies. On left panel we provide the percentage of patient who had a non-normal result on ADL subgroup scoresin the order Eyelid drop, Double Vision, Sit to Stand, Brush Teeth, Breathing, Swallowing, Chewing and Talking. On the right panel we provide the corresponding average score and standard deviation of the population.

Step 1: Let us first look systematically at the independence of each individual test score in MG-ADL, QMG, MG-CE and MG-QoL15. For MG-ADL and both the MGTX and BeatMG data set, we observe a statistically significant correlation (ρ >0.35, p<10^-10) within the group muscle 1 i.e. eye lid drop and diplopia, as well as within the group muscle 6 i.e. Chewing, Swallowing, Talking along with Teeth Brushing. The result is less clear in the ADAPT data set but for group muscle 1. However, we appreciate that the ADAPT data set (104 records) is small compared to the two others that have multiple time point with scores.

- For QMG and both the MGTX and BeatMG data set, we observed a strong correlation (ρ >0.83, p<10^-10) for the left arm score versus the right arm score, as well as left leg score with the right leg score. The left and right grip are correlated as well but with a lower correlation coefficient i.e. ρ >0.5, p<10^-10. We observe a statistically significant correlation (ρ >0.35, p<10^-10) within group muscle 1 for both data sets MGTX and BeatMG, and group muscle 6 only for the MGTX data set.
- Overall, the MGTX data set is the largest (2061 records) and gives higher correlation values than the Beat MG data set (728 records), for those individual test score that are correlated.
- We have the MG-Qol15 score for the ADAPT-MG data set only. As shown in Table 3, all individual test scores are statistically significantly correlated. We regroup individual test scores that are strongly corelated like left leg and right leg or left arm and right arm assessment in QMG into the same combine score. It is not feasible for the fatigue test score, without losing critical information on the patient state and therefore, we used a different technique.

Step 2: The fatigue score should be treated as a completely independent entity. Figure 3 shows the different pattern of fatigue obtained with 4 clusters that are clearly separated according to the signature function. We will present next an additional criterion to choose the optimum number of clusters beside cluster separation that is related to the construction of stochastic mapping with other clinical scores.

**Figure 3:**
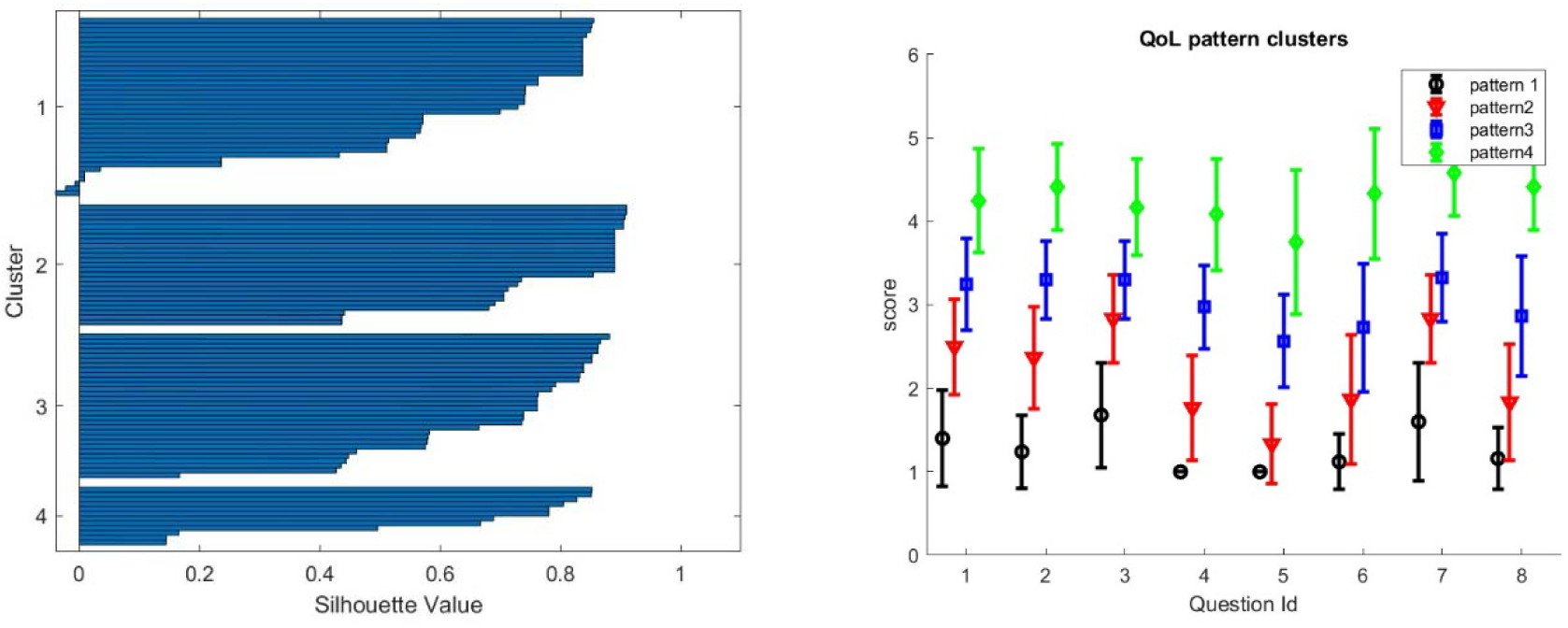
We obtain 4 well separated different patterns of QoL for the ADAPT-teleMG patients using a clustering algorithm. On the left panel, the signature silhouette shows the separation of clusters. On the right panel we show the 4 clusters with a monotinc progression on all invidual test score from low score to high score. The fact that the progression is monotonic across all questions is due to the strong correlation between individual test score as shown in Table 3.

Step 3: we review the stochastic maps we have been able to build across scores.

- One can map ADL and QMG component by component when there is clear association, i.e. same muscle group, like: ptosis, or diplopia, leg strength etc.… that are scored for both ADL and QMG. A similar analysis can be done for ADL and MG-CE with the ADAPT-MG data set. However, such maps would only be appropriate only if the correlation would be meaningful - see Figure 4. One can compare then the individual test scores of ADL and QMG only for ptosis, eyelid droop and talking. On the other hand, one can compare the ADL and MG-CE score only for ptosis, eyelid drop, limb and bulbar strength. We discard the cheek puff test from the MG-CE data set because of its poor accuracy as appreciated [12, 13].
- Figure 5 shows the the result for ptosis assessment by the clinician in QMG and eye lid drop as reported by the patient in the ADL score. From this graph one observes that only the most extreme score 0 and 3 are clearly in correspondence. One can appreciate also that the stochastic map from the MGTX and BeatMG data sets have similarities but are not identical. The variability of the intermediate scores are best described by the probability distribution. Figure 6 represents the stochastic map we obtained for double vision/diplopia. The direct map is close to a bijection, while the inverse map has the tendency to underscore the ADL value. Both stochastic maps are easier representations to interpret than the underlying row distribution of Fig A-2. Similar results can be obtained for the other available individual tests that have cross correlation value above 0.5 as noticed earlier – see Figure 4.

**Figure 4.**
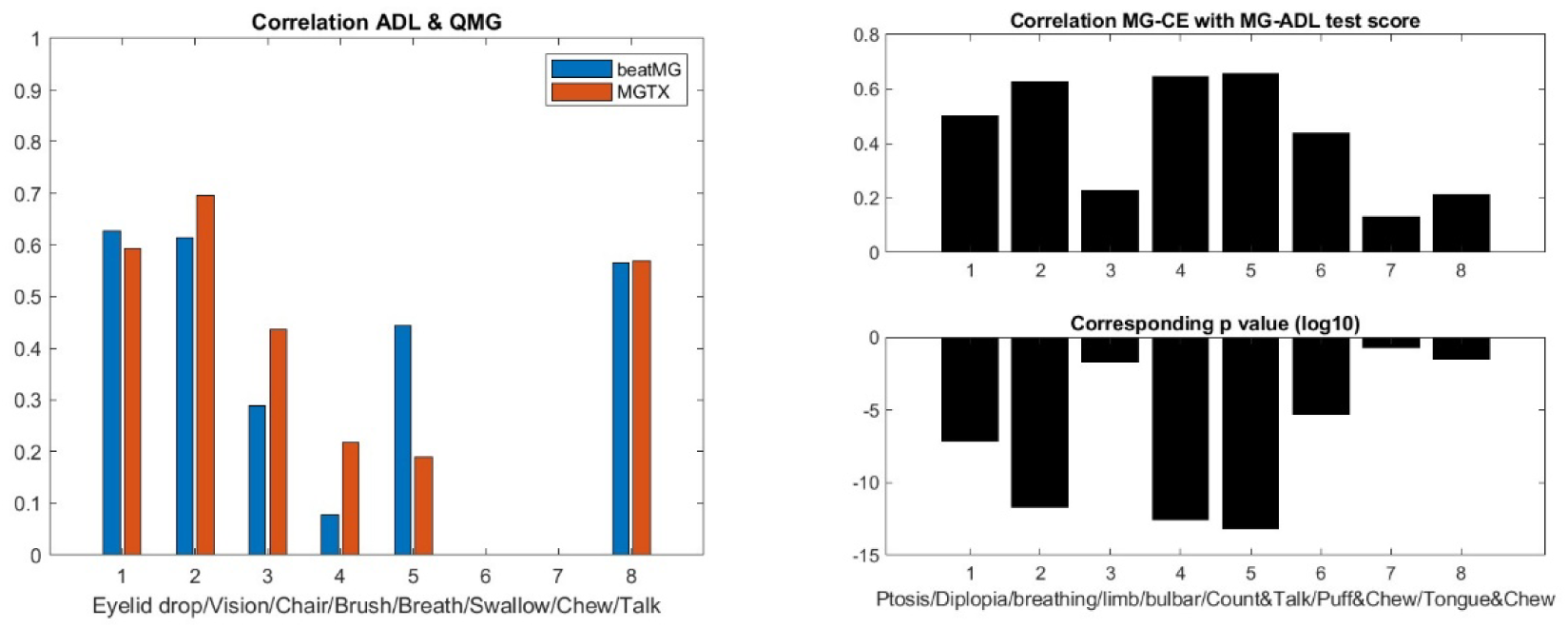
Correlation of the corresponding ADL and QMG score signs in the MGTX and BeatMG data set on the left panel (p-value are far less than 10^-20), and correlation of the corresponding MG-CE and MG-ADL scores in the ADAPT-MG study on the right panel.

**Figure 5:**
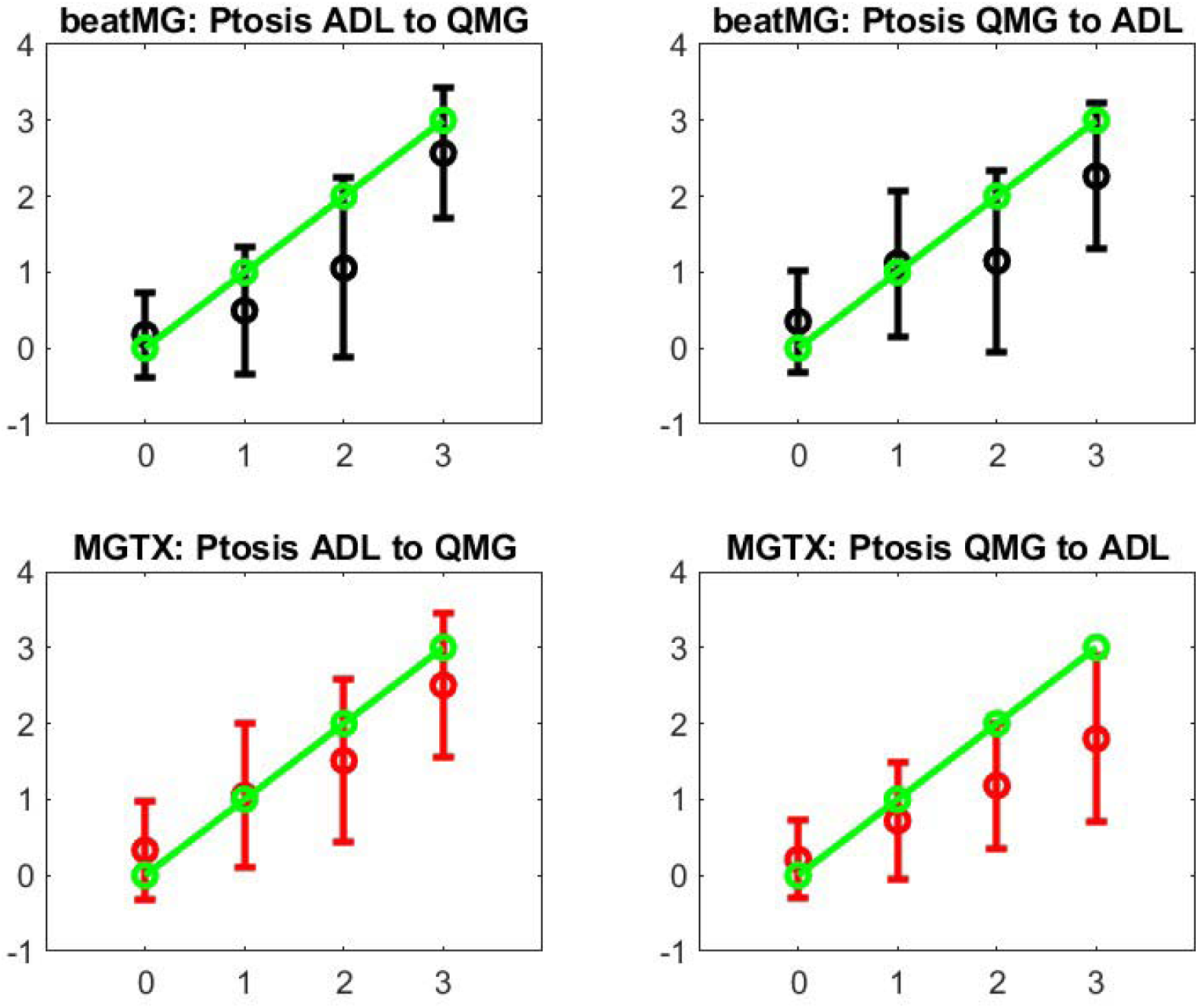
Stochastic Operators from ADL to QMG and its inverse. The green line represents what a perfect bijection should be. The left panel are for the direct map MGTX for the BeatMG and QMG data sets. The right panel is for the corresponding inverse map.

**Figure 6:**
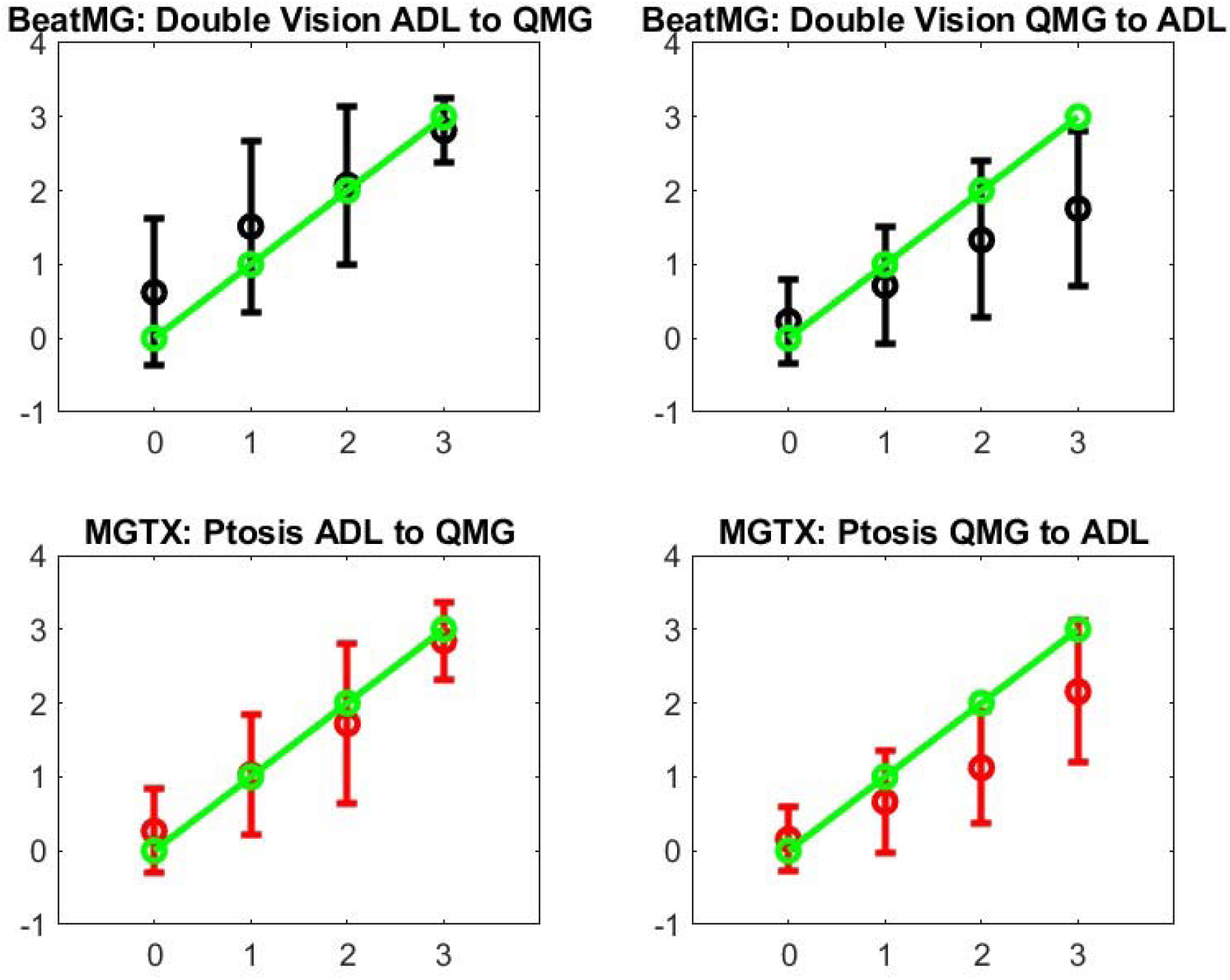
Stochastic Operators from ADL to QMG and its inverse for the double vision/diplopia test score. The green line represents what a perfect bijection should be. The left panel are for the direct map MGTX for the BeatMG and QMG data sets. The right panel is for the corresponding inverse map.

Step 4: Figure 7 shows the stochastic map between QoL and ADL. A similar map was obtained for QoL and QMG. Please notice that the mapping is nonlinear and is monotonic. It is convenient to get a monotonically increasing map: if we use more clusters in the classification of QoL patterns, the map is no longer monotonic because the stage overlaps too much. If we use less, we lower the overlap between the stochastic map values – see vertical bar of figure 7. While there is a moderate correlation between the ADL score and QoL score with ρ =0.61 and p=0.4 10^-12, Figure 4 gives us a stochastic map that is more structured than the global score correspondence one – see Figure A1 and take into account the correlation between individual QoL test score.

**Figure 7:**
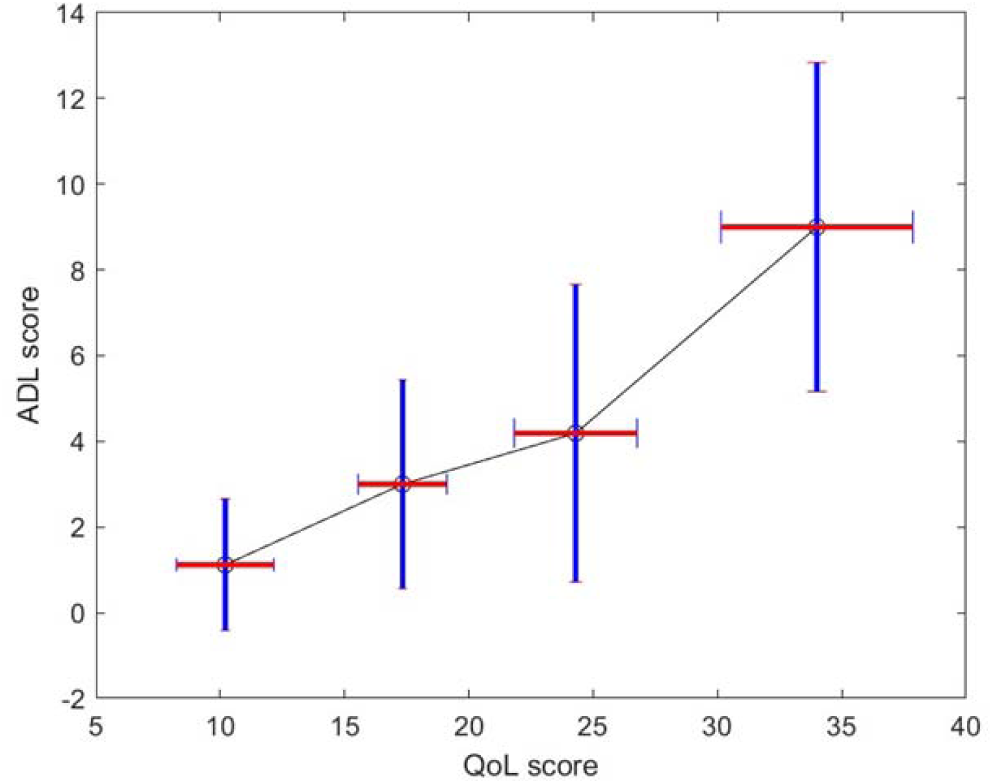
Average and standard deviation – vertical bar-of ADL score for each identified QoL patterns. The horizontal bar represents the mean and average QoL score foreach cluster.

Step 5:

Overall, we can complete a uniform score portrait of a patient with individual test score with ADL, QMG and QoL whenever a stochastic map is available along with its correspondent input has been collected.

We implement that strategy on the MGTX patient population, and derived for all patient the QoL score from the ADL one, based on the stochastic map statistical elements shown in Figure 7. ADL corresponds to patient report, and therefore appropriate to use such as the QoL as opposed to the QMG to generate another patient report score. More precisely, assuming a normal distribution, we could generate from the mean and standard deviation on the QoL score corresponding to each level 1 to 4 of the ADL score.

The score portrait can be optimized further, once we filter out from the score all the individual test score that have the most uncertainty or reliability like the cheek puff in MG-CE, or use weighted combination that increase the predictability of patient trajectory as defined previously [19].

For now, we will use the global (ADL,QMG,QoL) triplet as the score portrait as a simplification and concentrate on the MGTX data set that had the best success in providing new results.

To generate a virtual population from that data set of score portrait (ADL,QMG,QoL) for each patient of the MGTX clinical study, we used a standard Montecarlo method. We assume that the uncertainty on the ADL and QMG score is given by a normal distribution of standard deviation being one which seems to be a conservative estimate [13]. Using a scaling factor of 10 with the Montecarlo method, the virtual population has about 1110 patients.

Following the clustering technique described in [16], we compute the mean trajectories of these virtual patients with 4 clusters and keep a good separation between clusters as shown in the silhouette portrait as shown in Figure A3. This new result seems to bring additional fine details that were not available with the original MGTX data set: we found then that 3 clusters was the largest number allowed according to the signature function.

We can generate a virtual population with QoL pattern score for the MGTX patients that do not have QoL records. Figure 8 shows the main trajectories for the QMG and QoL score. For completeness the ADL score trajectories are shown in Figure A4 of the appendix.

**Figure 8.**
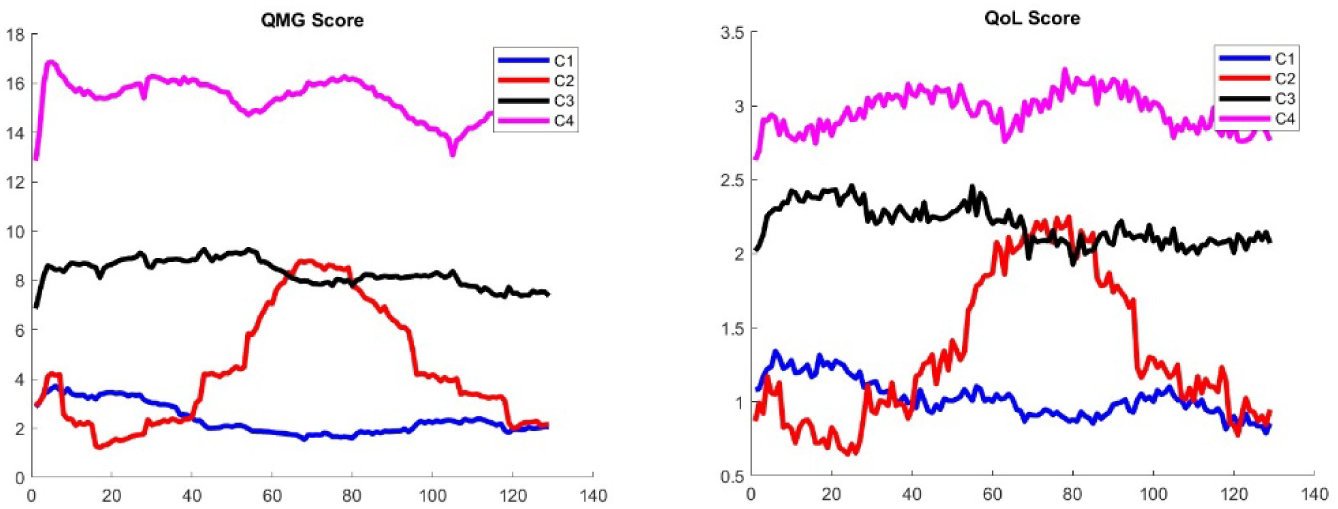
Classification of Trajectories for the QMG (left panel) and QoL score (right panel) of the virtual population generated from the MGTX patients. The QoL score that has been generated from the ADL score using the stochastic map of Figure 7.

It should be appreciated that the clustering was applied to the score portrait including all three scores at once, instead of each individual score. It is then remarkable that the ranking of clusters from greatest improvement– cluster one – to treatment resistant– cluster 4, matches the same order for all three scores. While the virtual population offers a much larger sample to cluster, it is not clear why clustering should work better than the original MGTX set. However, the following results provides some validation of the results:

Figure A5 in appendix shows the benefit of thymectomy for patient outcome expressed as daily prednisone dose. As expected, the thymectomy patients were more commonly in Cluster 1, which identified the subjects who responded best to treatment. Figure A6 graphics confirms that the patients who do best have the lowest number of doses of azathioprine, and conversely the treatment-resistant patients had highest use of azathioprine

In addition, we observe a new phenomenon with cluster 2 that represents only 7 % of the total virtual population. For those patients, the mean trajectory differs significantly from the three others that stay relatively flat. We observe that Cluster 2 has a significantly higher percentage of patient older than 50 – see Figure 9 left panel. In our previous study with the MGTX data set itself, i.e. not the virtual population using the score portrait generated, those individuals were included in cluster one that was the largest.

**Figure 9.**
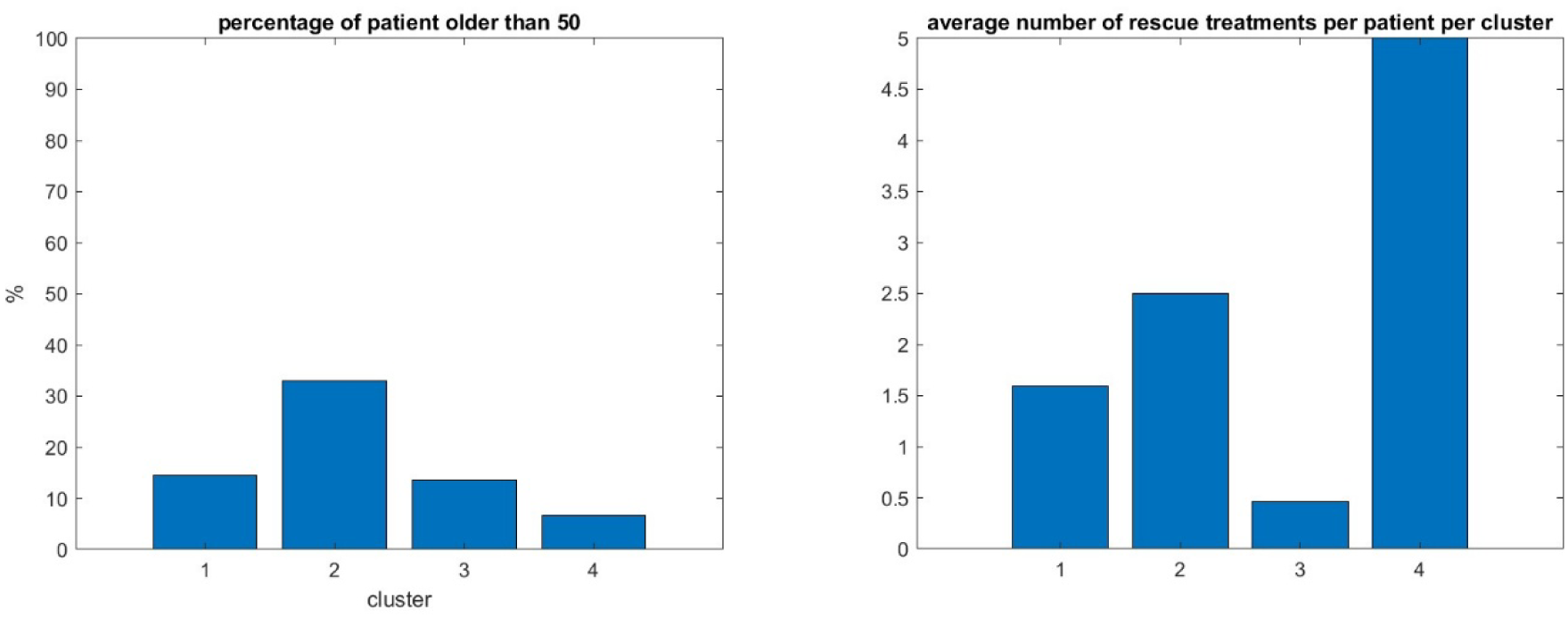
percentage of patient over 50 in the same four clusters (left panel), number of rescue therapy session per patient per cluster (right panel)

We hypothesize that the benefit of generating a virtual population with a score portrait that includes all three scores is a finer granularity that can identify this kind of singular behavior. It remains to be seen if the daily drug dosage could have been managed differently for those patients. The distribution of patients over the four clusters of this virtual population who went through rescue therapy i.e. plasmapheresis and Intravenous immunoglobulin, confirms that cluster four that corresponds to patient with the less successful therapy has indeed the greatest use of rescue therapy – see Figure 9 right panel. But since the proportion of patient having rescue therapy is relatively small i.e. 14 %, it will take a larger cohort to better understand the factors producing these clusters.

## Discussion

We have successfully developed a step-by-step method to create a score portrait for patients with MG. This portrait integrates all available information from the QMG, MG-ADL, and MG-QoL scores, drawing from a representative set of independent clinical trial datasets. A key challenge was managing studies with incomplete records or missing scores. The trade-off for this comprehensive construction is that the score portrait is no longer a single number, but rather a probability distribution. The primary advantage of this approach is its ability to generate a uniform metric across various clinical studies, even those not initially designed with matching protocols. However, this comes with a level of uncertainty embedded in the score portrait, which reflects the inherent limitations when information is missing. This concept is broadly applicable to diverse clinical studies beyond just MG.While this feasibility study employed relatively simple statistical approaches, such as stochastic maps between score types or subsets of score types, it clearly demonstrated that none of the individual scores are ideal for instance, correlations between QoL test values can be strong. To some extent, correlations with muscle groups in QMG, ADL, or MG-CE are non-negligible. To define a more robust score that effectively combines physical examination and patient outcomes—as our ADL, QMG, and QoL combination aims to do—further clinical guidance and validation are essential to accurately predict patient trajectories [24, 25]. Generating virtual populations of digital twin patients from numerous independent clinical trials appears to be a promising strategy to address the challenge of MG’s rare disease status, characterized by many clinical trials but few patients. The practical value of these virtual populations has yet to be demonstrated robustly [21]. However, our preliminary results, built on the rigorously performed and complex MGTX study, suggest that exploring virtual populations can reveal finer details on patient trajectories within small subgroups. These insights might otherwise be missed in standard analyses applied to clinical trials. We intentionally kept our algorithmic approach, from step 1 to 5, systematic. This provides an opportunity to re-evaluate current standards for scores and minimum manifestations. Nevertheless, gaps likely exist in clinical study protocols, particularly concerning the impact of patient personality traits that impact patient reported outcome measures, for example openness, resilience and adaptability to disability. These are is a crucial components of how patients cope with the disease and interact with clinicians during evaluations.

### Conclusion

This study successfully developed a novel “Myasthenia Gravis Portrait” that integrates various disease severity scores (QMG, ADL, QoL) into a unified, probability-based metric. This approach addresses the challenge of diverse scoring systems and incomplete data across different clinical trials, enabling a more standardized assessment of disease progression. The creation of this portrait facilitates the generation of virtual patient populations, a crucial step for applying machine learning techniques to better understand and predict individual patient trajectories in this rare disease. While further clinical validation and the incorporation of additional patient assessment factors like psychological state and objective fatigue measures are needed, this framework offers a promising path toward more robust and comprehensive MG research.

## Data Availability

All deidentified data used in this study are available from the authors upon reasonable request.

---

**Table 3:**
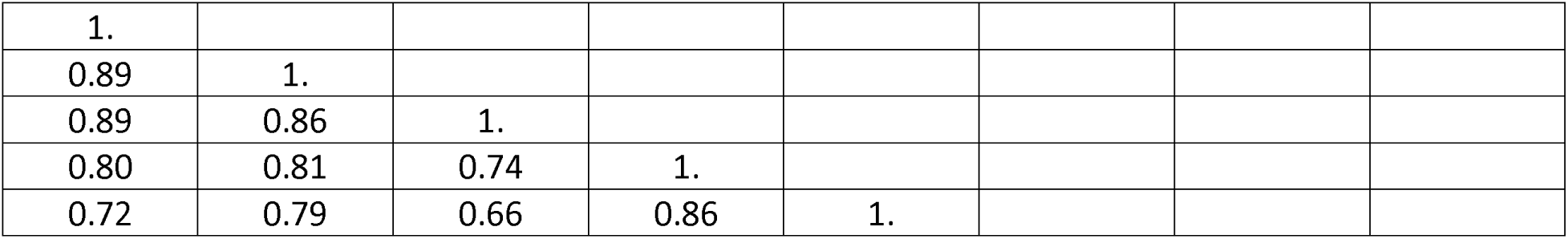
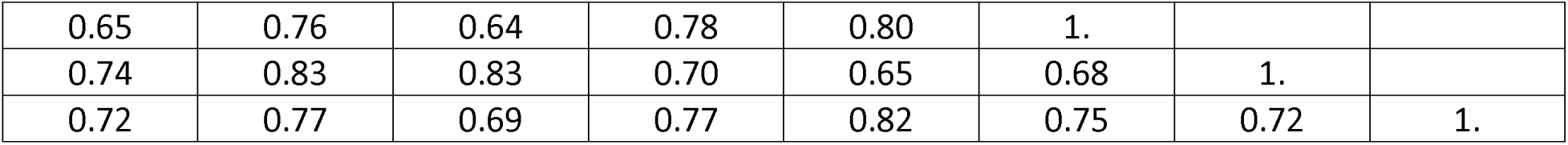
autocorrelation of QOL score in the MGnet study with p<0.0000000000001.

|  |  |  |  |  |
| --- | --- | --- | --- | --- |
| 1. |  |  |  |  |
| 0.89 | 1. |  |  |  |
| 0.89 | 0.86 | 1. |  |  |
| 0.80 | 0.81 | 0.74 | 1. |  |
| 0.72 | 0.79 | 0.66 | 0.86 | 1. |

|  |  |  |  |  |  |  |  |
| --- | --- | --- | --- | --- | --- | --- | --- |
| 0.65 | 0.76 | 0.64 | 0.78 | 0.80 | 1. |  |  |
| 0.74 | 0.83 | 0.83 | 0.70 | 0.65 | 0.68 | 1. |  |
| 0.72 | 0.77 | 0.69 | 0.77 | 0.82 | 0.75 | 0.72 | 1. |

## APPENDIX

**Figure A-1.**
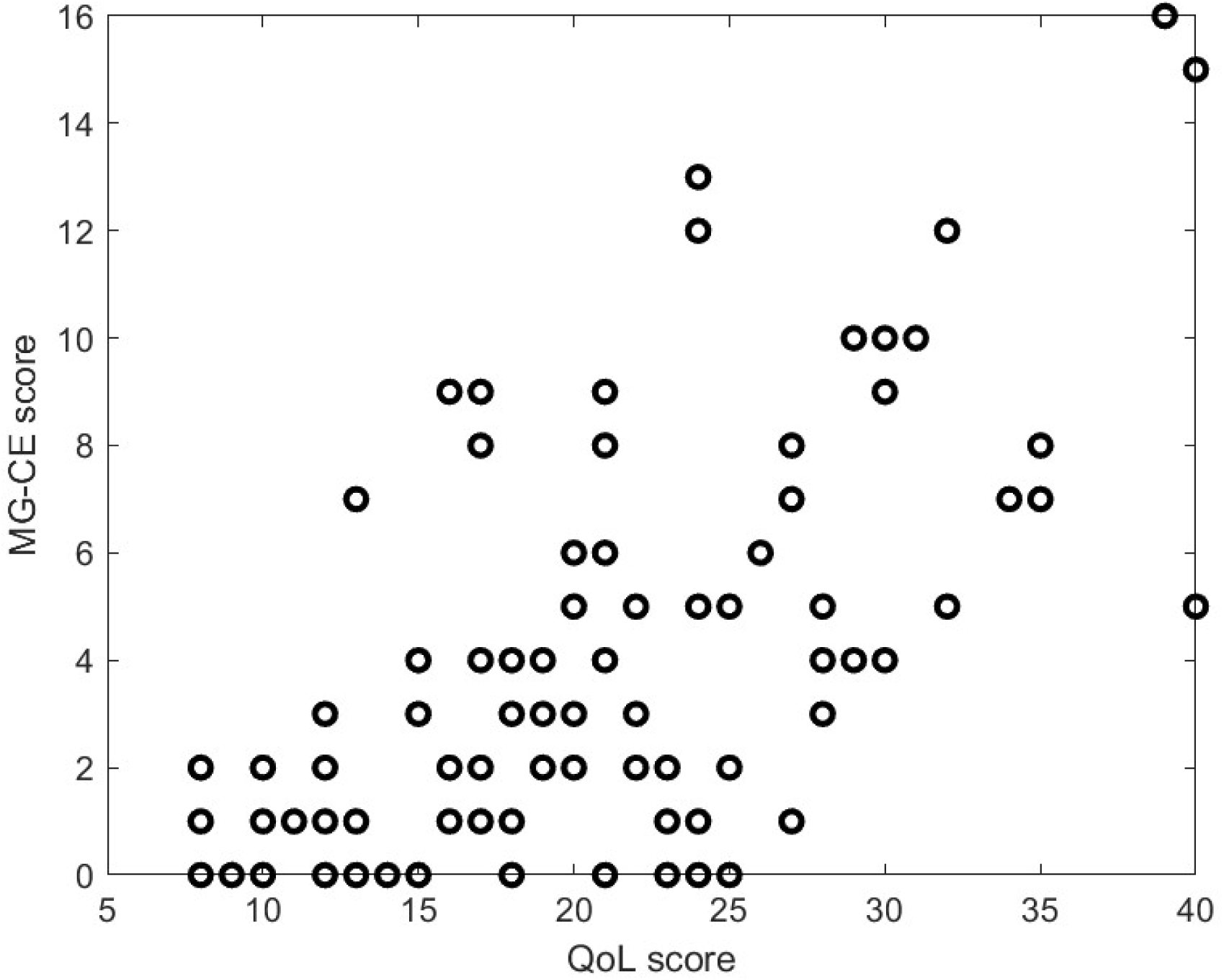
Global MG-ADL score versus QOL score

**Figure A-2:**
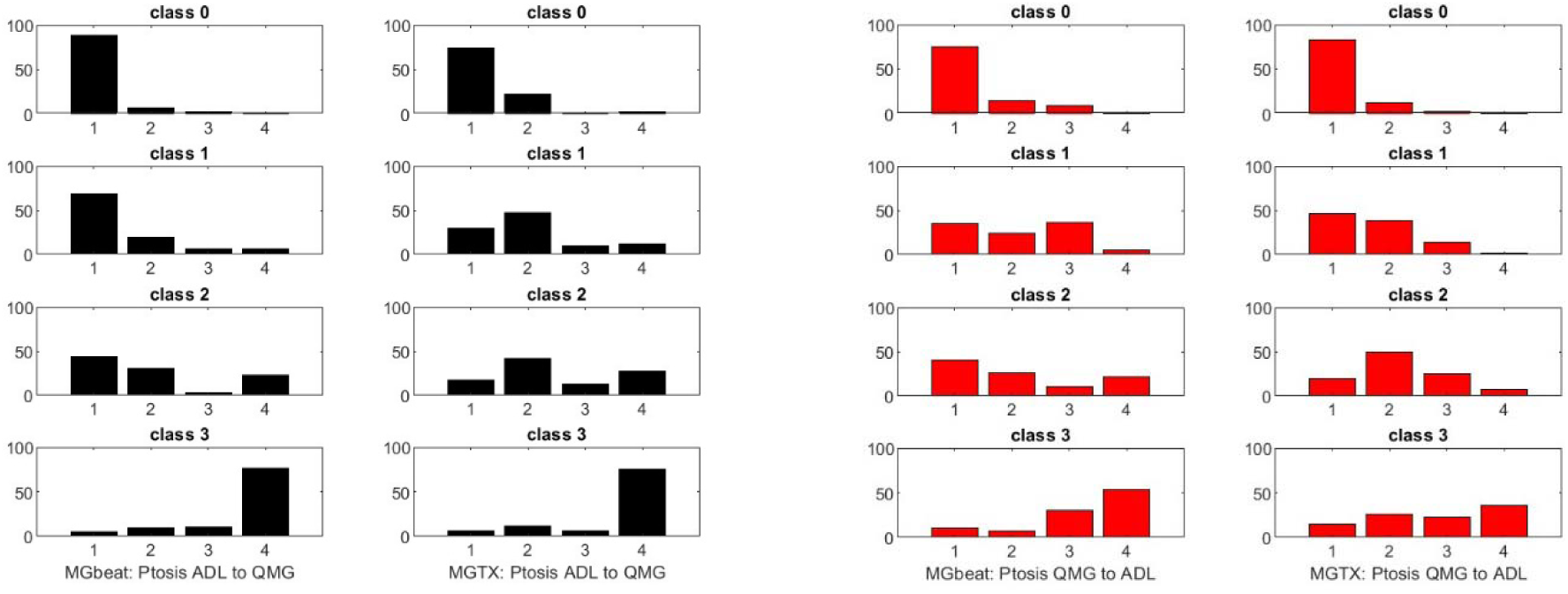
Considering two score A and B, one can compute the percentage Distribution of Score sign in B from a given Score in A see left panel or vice versa – see right panel. In this graphic A and B are respectively the ADL and QMG score

**Figure A-3.**
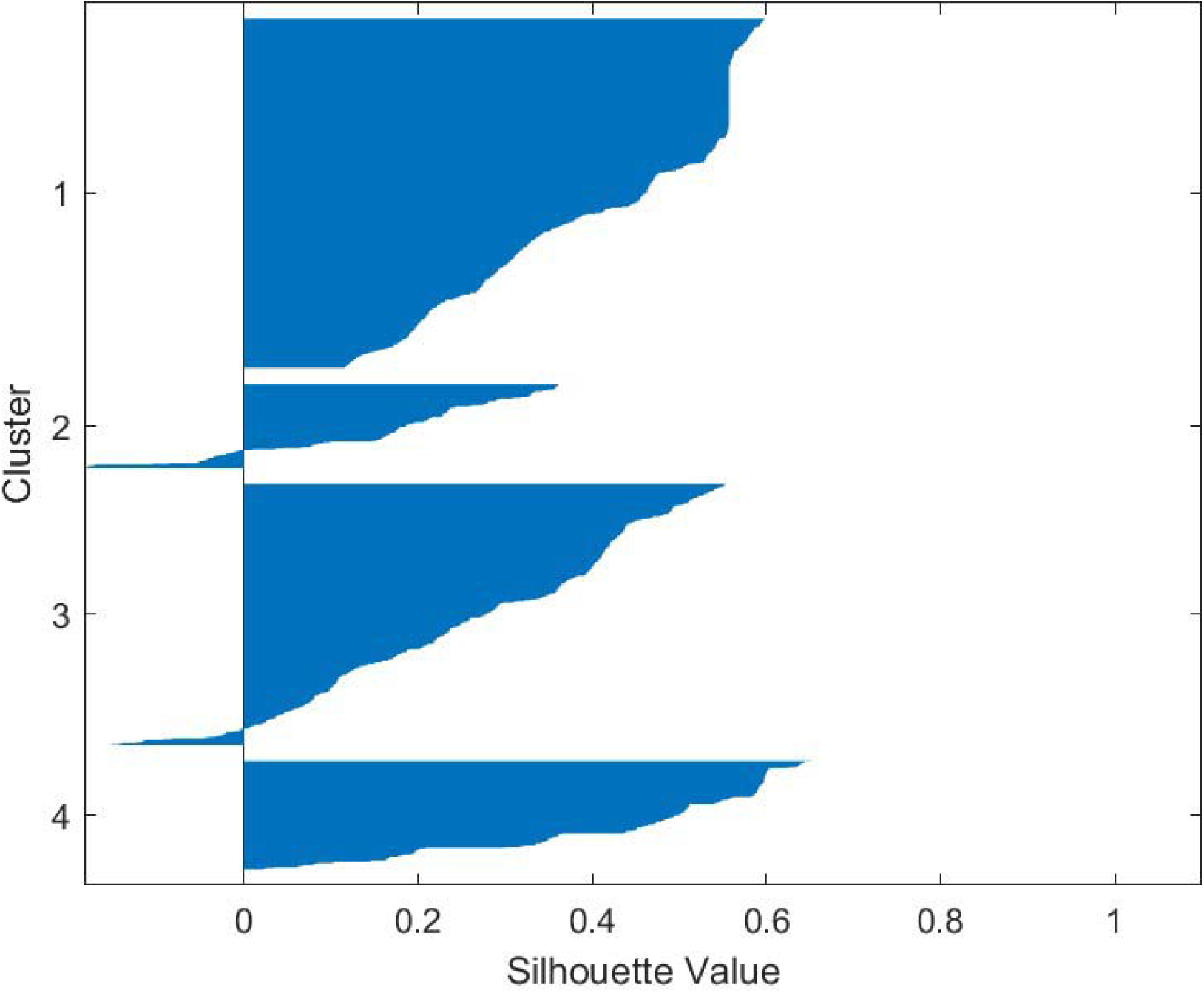
Silhouette map for the clustering of the score portrait of the virtual population of 1110 individuals.

**Figure A-4.**
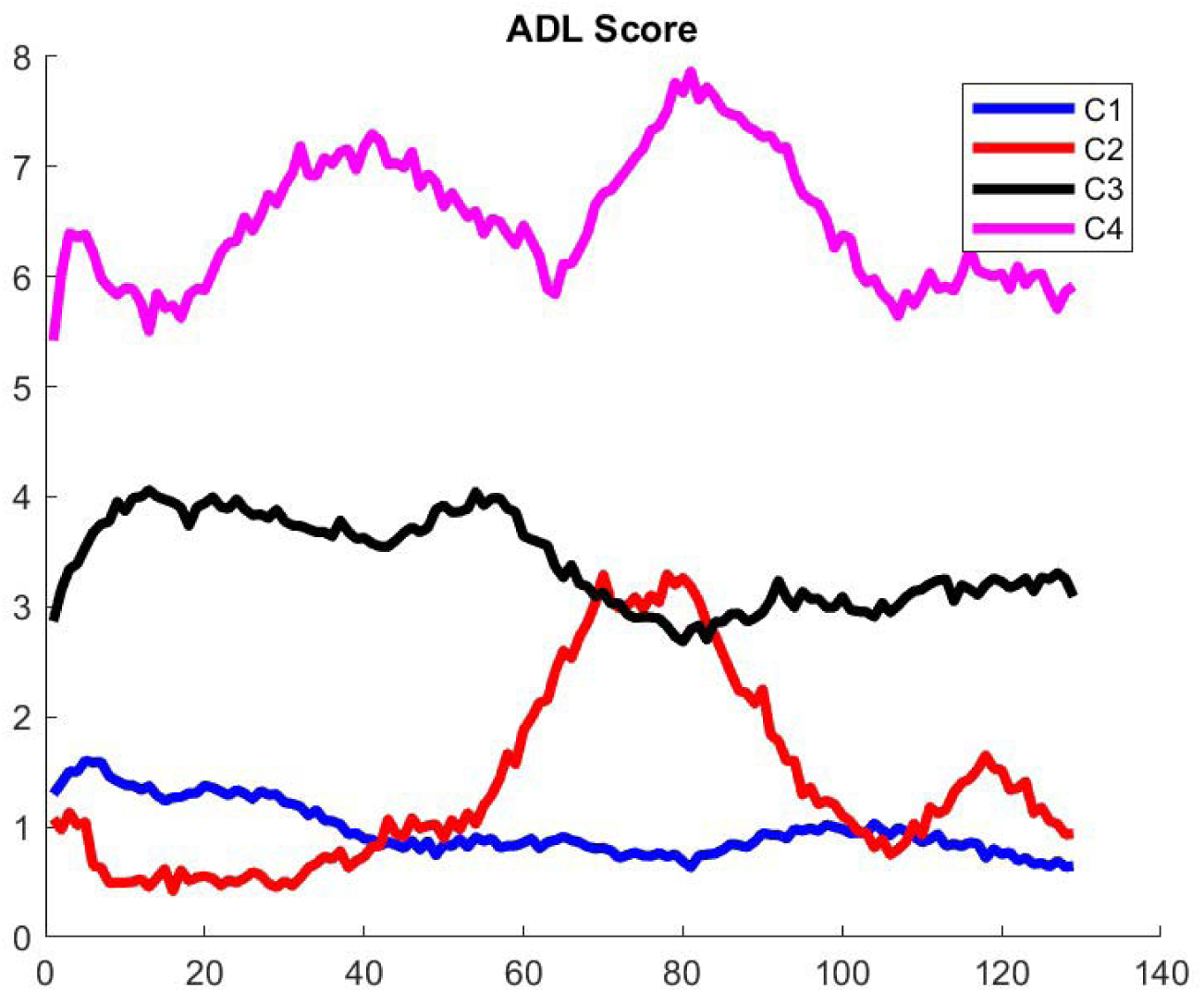
Classification of Trajectories for the ADL score of the virtual population generated from the MGTX patient. Figure 8 and A-3 provide all together the completed score portrait of these patients.

**Figure A5.**
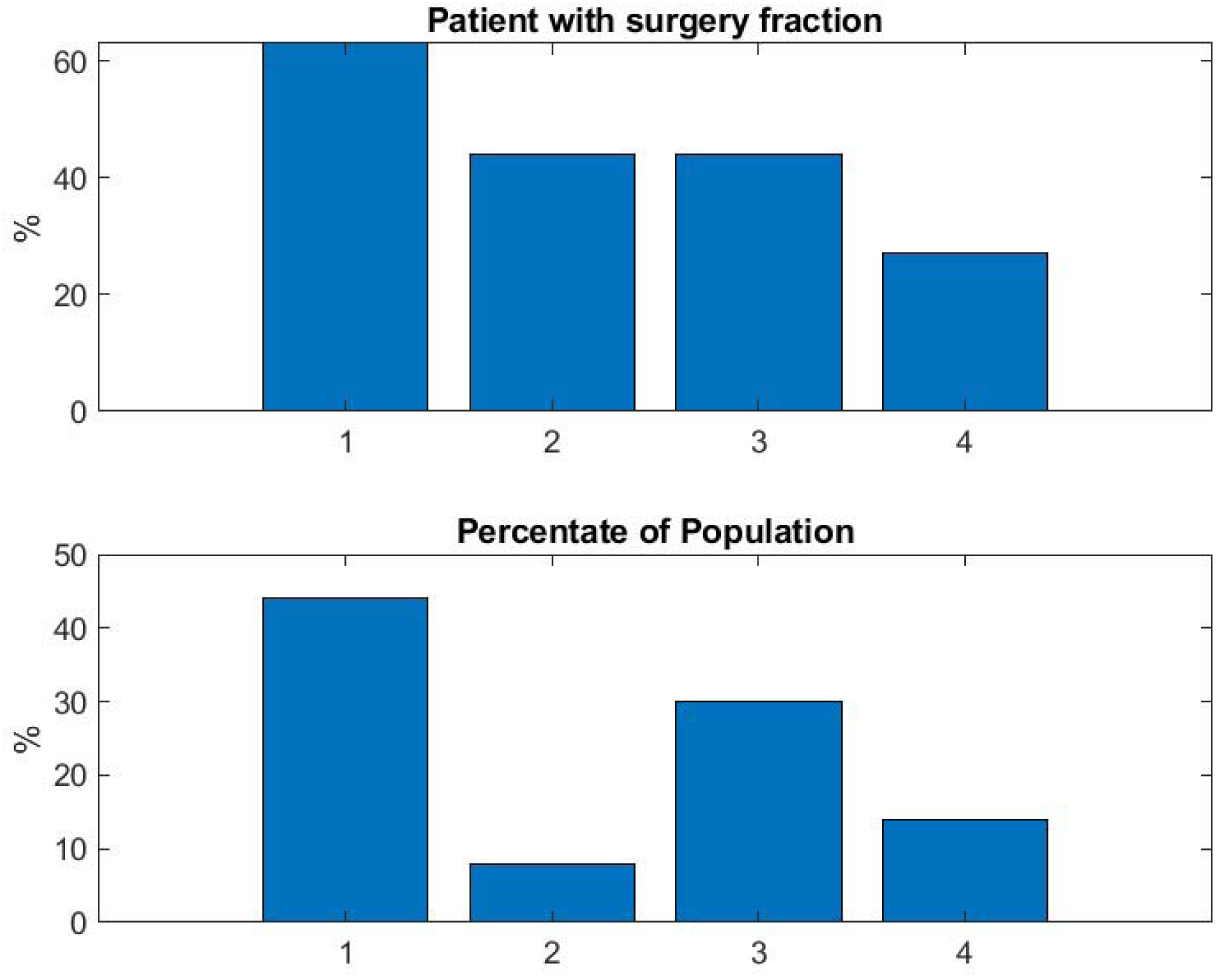
Result confirms the benefit of thymectomy for patient outcome expressed as daily prednisone dose. The thymectomy patients were more commonly in Cluster 1, which identified the subjects who responded best to treatment.

**Figure A6.**
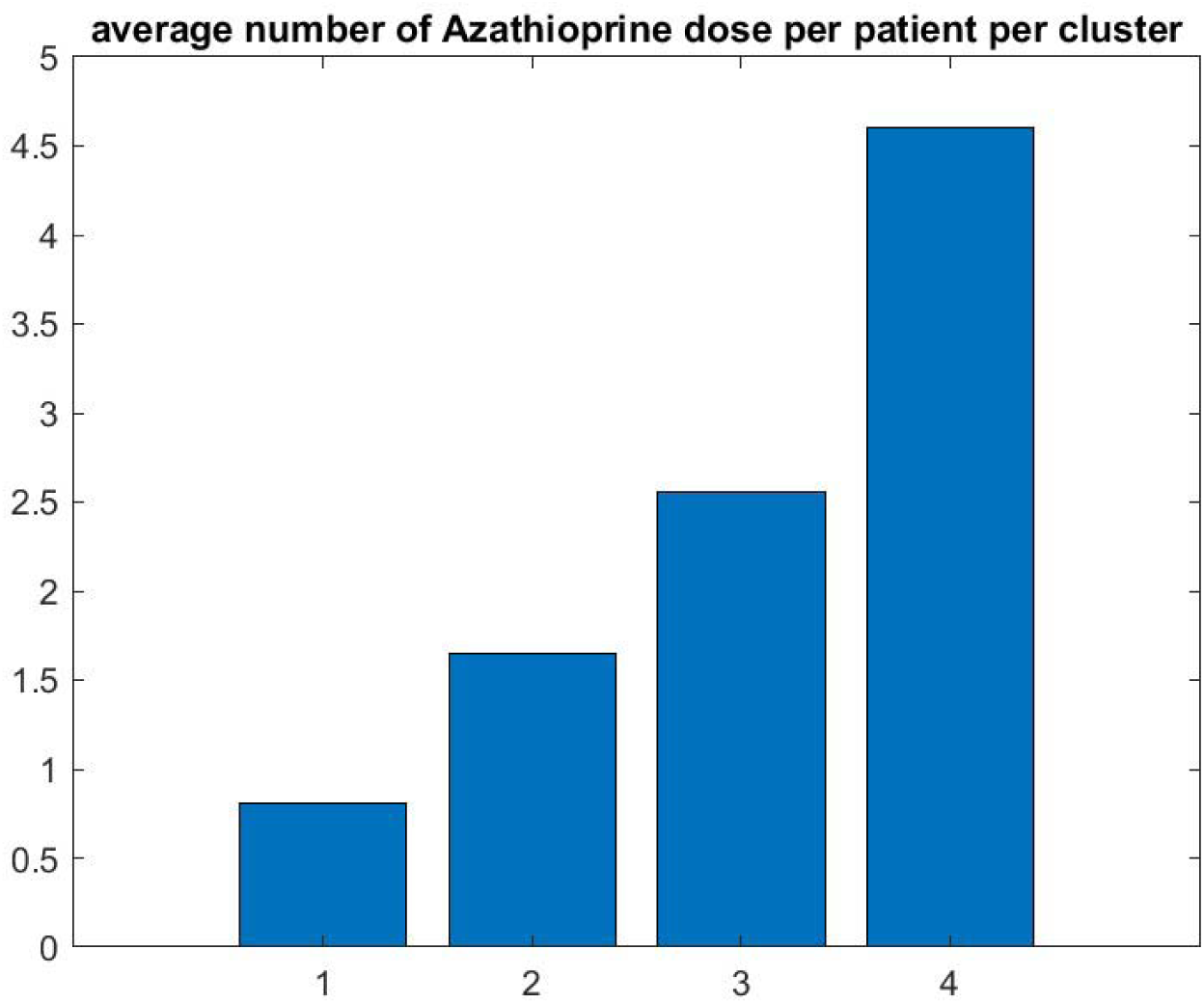
Graphic confirms that the patients who do best (respectively worse) have the lowest (respectively highest) number of doses of azathioprine

## References

1. Kaminski, H.J., Sikorski, P., Coronel, S.I. & Kusner, L.L. Myasthenia gravis: the future is here. J. Clin. Invest. 134, e179742 (2024). 10.1172/JCI179742

2. Jaretzki A 3rd, Barohn RJ, Ernstoff RM, Kaminski HJ, Keesey JC, Penn AS, Sanders DB. Myasthenia gravis: recommendations for clinical research standards. Task Force of the Medical Scientific Advisory Board of the Myasthenia Gravis Foundation of America. Neurology. 2000 Jul 12;55(1):16–23.

3. Barohn RJ, McIntire D, Herbelin L, Wolfe GI, Nations S, Bryan WW. Reliability testing of the quantitative myasthenia gravis score. Ann N Y Acad Sci. 1998 May 13;841:769–72. doi: 10.1111/j.1749-6632.1998.tb11015.x. PMID: 9668327.

4. Burns, T.M., Conaway, M.R., Cutter, G.R. & Sanders, D.B. Construction of an efficient evaluative instrument for myasthenia gravis: the MG composite. Muscle Nerve 38, 1553–1562 (2008).

5. Burns TM, Conaway M, Sanders DB; MG Composite and MG-QOL15 Study Group. The MG Composite: A valid and reliable outcome measure for myasthenia gravis. Neurology. 2010 May 4;74(18):1434–40. doi: 10.1212/WNL.0b013e3181dc1b1e. PMID: 20439845; PMCID: PMC3462556.

6. Janssen, M.F., Dewilde, S., Wolfe, G.I. & Muppidi, S. Psychometric properties of MG-ADL items and MG-ADL score: an assessment of distributional characteristics, validity and factor structure in two large datasets. J. Neurol. Sci. 463, 123135 (2024). 10.1016/j.jns.2024.123135

7. Muppidi, S. et al. Utilization of MG-ADL in myasthenia gravis clinical research and care. Muscle Nerve 65, 630–639 (2022).

8. Annabel M. Ruiter, Jan J.G.M. Verschuuren, Martijn R. Tannemaat, Fatigue in patients with myasthenia gravis. A systematic review of the literature, Neuromuscular Disorders 30 (2020) 631–639 Review www.elsevier.com/locate/nmd

9. T.M. Burns, R. Sadjadi, K. Utsugisawa, et al. International clinicometric evaluation of the MG-QOL15, resulting in slight revision and subsequent validation of the MG-QOL15r, Muscle Nerve, 54 (6) (2016), pp. 1015–1022

10. Guidon, A.C. et al. Telemedicine visits in myasthenia gravis: expert guidance and the Myasthenia Gravis Core Exam (MG-CE). Muscle Nerve 64, 270–276 (2021). 10.1002/mus.27260

11. S. Dewilde, M.F. Janssen, N. Tollenaar, et al., Concordance between patient- and physician-reported MG-ADL scores, Muscle Nerve, 68 (2023), pp. 65–72

12. Marc Garbey, Quentin Lesport, Helen Girma, Gül⍰en Öztosen, Mohammed Abu-Rub, Amanda C. Guidon, Vern Juel, Richard Nowak, Betty Soliven, Inmaculada Aban, Henry J. Kaminski, Application of Digital Tools and Artificial Intelligence to the Myasthenia Gravis Core Examination, Frontiers in Neurology, Vol 15, 2024 URL=https://www.frontiersin.org/journals/neurology/articles/10.3389/fneur.2024.1474884

13. Marc Garbey, Quentin Lesport, Helen Girma, Gülşen Öztosun, Henry J. Kaminski, A Quantitative Study of Factors Influencing Myasthenia Gravis Telehealth Examination Score https://www.medrxiv.org/content/10.1101/2024.07.24.24310934v1, Muscle and Nerve, in press.

14. Wolfe, G.I., Kaminski, H.J., Jaretzki, A. 3rd, Swan, A. & Newsom-Davis, J. Development of a thymectomy trial in nonthymomatous myasthenia gravis patients receiving immunosuppressive therapy. Ann. N. Y. Acad. Sci. 998, 473–480 (2003). 10.1196/annals.1254.061

15. Wolfe, G.I. et al. Randomized trial of thymectomy in myasthenia gravis. N. Engl. J. Med. 375, 511–522 (2016).

16. Nowak, R. J., et al., Phase 2 Trial of Rituximab in Acetylcholine Receptor Antibody-Positive Generalized Myasthenia Gravis: The BeatMG Study, Muscle & Nerve (2021).

17. Vallée A. Digital twin for healthcare systems. Front Digit Health. 2023 Sep 7;5:1253050. doi: 10.3389/fdgth.2023.1253050. PMID: 37744683; PMCID: PMC10513171.

18. Mathieu F. Janssen, Sarah Dewilde, Gil I. Wolfe, Srikanth Muppidi, Glenn Phillips, Psychometric properties of MG-ADL items and MG-ADL score: An assessment of distributional characteristics, validity and factor structure in two large datasets, Journal of the Neurological Sciences, Volume 463, 2024, 10.1016/j.jns.2024.123135.

19. Marc Garbey, Quentin Lesport, Henry J. Kaminski, Construction of patient trajectories to model clinical trial outcomes: Application to Myasthenia Gravis, medrxiv preprint series, https://www.medrxiv.org/content/10.1101/2025.04.11.25325663v1

20. Teutonico D, Musuamba F, Maas HJ, Facius A, Yang S, Danhof M, Della Pasqua O. Generating Virtual Patients by Multivariate and Discrete Re-Sampling Techniques. Pharm Res. 2015 Oct;32(10):3228–37. doi: 10.1007/s11095-015-1699-x. Epub 2015 May 21. PMID: 25994981; PMCID: PMC4577546.

21. Guo Y, Guo T, Knibbe CAJ, Zwep LB, van Hasselt JGC. Generation of realistic virtual adult populations using a model-based copula approach. J Pharmacokinet Pharmacodyn. 2024 Dec;51(6):735–746. doi: 10.1007/s10928-024-09929-4. Epub 2024 Jun 6. PMID: 38844624; PMCID: PMC11579194.

22. Gold R, Schneider-Gold C. Current and future standards in treatment of myasthenia gravis. Neurotherapeutics. 2008 Oct;5(4):535–41. doi: 10.1016/j.nurt.2008.08.011. PMID: 19019304; PMCID: PMC4514693.

23. Lehnerer Sophie, Stegherr Regina, Grittner Ulrike, Stein Maike, Gerischer Lea, Stascheit Frauke, Herdick Meret, Doksani Paolo, Meisel Andreas, Hoffmann Sarah, The burden of disease in seronegative myasthenia gravis: a patient-centered perspective, Frontiers in Immunology, Vol 16, 2025, https://www.frontiersin.org/journals/immunology/articles/10.3389/fimmu.2025.1555075

24. Meisel, A., Saccà, F., Spillane, J., Vissing, J. & MG Collegium Sub-committee. Expert consensus recommendations for improving and standardising the assessment of patients with generalised myasthenia gravis. Eur. J. Neurol. 31, e16280 (2024). 10.1111/ene.16280

25. Watanabe, G. et al. Cutoffs on severity metrics for minimal manifestations or better status in patients with generalized myasthenia gravis. Front. Immunol. 15, 1502721 (2024). 10.3389/fimmu.2024.1502721C4793785.

26. Bjelland I, Dahl AA, Haug TT, Neckelmann D. The validity of the Hospital Anxiety and Depression Scale. An updated literature review. J Psychosom Res. 2002 Feb;52(2):69–77. doi: 10.1016/s0022-3999(01)00296-3. PMID: 11832252.

